# Left atrial strain in multisystem inflammatory syndrome in children (MIS-C) and associations with systemic inflammation and cardiac injury

**DOI:** 10.1101/2023.05.22.23290346

**Authors:** Bryan M. Jepson, Matthew Beaver, John L. Colquitt, Dongngan T. Truong, Hillary Crandall, Carol McFarland, Richard Williams, Zhining Ou, Devri Jensen, L. LuAnn Minich, Edem Binka

## Abstract

**Background:** Multisystem inflammatory syndrome in children (MIS-C) commonly involves cardiac injury with both systolic and diastolic dysfunction. Left atrial strain (LAS) detects subclinical diastolic dysfunction in adults but is infrequently used in children. We evaluated LAS in MIS-C and the associations with systemic inflammation and cardiac injury.

**Methods:** In this retrospective cohort study, conventional parameters and LAS (reservoir [LAS-r], conduit [LAS-cd], and contractile [LAS-ct]) obtained from admission echocardiograms of MIS-C patients were compared to healthy controls and between MIS-C patients with and without cardiac injury (BNP >500 pg/ml or troponin-I >0.04 ng/ml). Correlation and logistic regression analyses were performed to assess LAS associations with admission inflammatory and cardiac biomarkers. Reliability testing was performed.

**Results:** Median LAS components were reduced in MIS-C patients (n=118) compared to controls (n=20) (LAS-r: 31.8 vs. 43.1%, p<0.001; LAS-cd: -28.8 vs. -34.5%, p=0.006; LAS-ct: -5.2 vs. -9.3%, p<0.001) and reduced in MIS-C patients with cardiac injury (n=59) compared to no injury (n=59) (LAS-r: 29.6 vs. 35.8%, p=0.001; LAS-cd: -26.5 vs. -30.4%, p=0.036; LAS-ct: -4.6 vs. -9.3%, p=0.008). An LAS-ct peak was absent in 65 (55%) MIS-C patients but present in all controls (p<0.001). Procalcitonin had strong correlation with averaged E/e’ (r=0.55, p=0.001); ESR had moderate correlation with LAS-ct (r=-0.41, p=0.007); BNP had moderate correlation with LAS-r (r=-0.39, p<0.001) and LAS-ct (r=0.31, p=0.023), and troponin-I had only weak correlations. No strain indices were independently associated with cardiac injury on regression analysis. Intra-rater reliability was good for all LAS components; and inter-rater reliability was good to excellent for LAS-r, and fair for LAS-cd and LAS-ct.

**Conclusions:** LAS analysis, particularly the absence of a LAS-ct peak, was reproducible and may be superior to conventional echocardiographic parameters for detecting diastolic dysfunction in MIS-C. No strain parameters on admission were independently associated with cardiac injury.

## Introduction

Children with severe acute respiratory syndrome coronavirus 2 (SARS-CoV-2) infection may develop a post-infectious, hyper-inflammatory condition termed multisystem inflammatory syndrome in children (MIS-C).^1-5^ While MIS-C has been reported to affect all organ systems, up to 80% of children with MIS-C have cardiovascular involvement ranging in severity from a well-compensated state to refractory shock at the time of presentation.^2,3,6-10^ Strain echocardiography has been increasingly used in the MIS-C population^11-15^ and abnormalities in left ventricular (LV) strain at presentation persist through short term follow up, even in children with normal LV ejection fraction (LVEF).^16-19^ Less is known about the use of left atrial strain (LAS) to assess diastolic dysfunction in children with MIS-C.

LAS is used in adults as a surrogate measure of LV diastolic dysfunction^20-25^ and has been increasingly investigated in children with MIS-C. Matsubara *et al*. reported that peak LAS had the strongest association with cardiac injury and was superior to ventricular deformational parameters.^16^ Compared to traditional echocardiographic parameters, phasic LAS analysis has the potential to discern subclinical diastolic dysfunction,^20,21,26^ but which LAS components may be the best predictor of diastolic dysfunction remains unclear. The strength of the association between phasic LAS components and cardiac injury warrants further investigation in children with MIS-C.

The aims of this study were to 1) evaluate phasic LAS components (reservoir [LAS-r], conduit [LAS-cd], and contractile [LAS-ct]) in children presenting with MIS-C, 2) determine the association of LAS with laboratory markers of inflammation and cardiac injury upon admission in MIS-C, and 3) assess the reproducibility of phasic LAS analysis in this population.

## Methods

### Study Population

This retrospective, single-center, cohort study included patients age <19 years hospitalized with MIS-C between April 1, 2020 and December 31, 2021. Patients with pre-existing cardiomyopathy, congenital heart disease, or prior exposure to cardiotoxic therapies were excluded. MIS-C was diagnosed using the case definition outlined by the U.S. Centers for Disease Control and Prevention.^27^ Demographic and clinical data were collected and managed using REDCap electronic data capture tools hosted at our institution.^28,29^ Clinical data included admission laboratory values (C-reactive protein [CRP], D-dimer, erythrocyte sedimentation rate [ESR], ferritin, procalcitonin, brain natriuretic peptide [BNP], and troponin-I), intensive care unit (ICU) admission, extracorporeal membrane oxygenation (ECMO) support, and maximum level of respiratory support. Cardiac injury was defined as either an admission BNP >500 pg/ml^16^ or troponin-I >0.04 ng/ml.

In the absence of universally accepted reference ranges for LAS components in children, we identified 20 healthy children who had a normal echocardiogram for evaluation of chest pain or murmur during the study period to form the control group. To obtain representative controls, the study cohort was divided into quartiles by age and the distribution of sex was determined for each quartile. Control cases were then selected at random from the study period to match the sex distribution within each age quartile.

### Echocardiography

MIS-C study patients underwent two-dimensional and Doppler echocardiography at the time of admission as part of routine clinical care and studies were reviewed retrospectively.

Echocardiograms were performed using either a Philips iE33 or Philips EPIQ CVx ultrasound machine (Philips Medical Systems, Massachusetts) for both study patients and healthy controls. Standard echocardiographic measurements were obtained in accordance with American Society of Echocardiography guidelines.^30,31^ If echocardiogram reports did not record values for the pre-designated study variables, measurements were made retrospectively. Echocardiographic values collected included qualitative valvar regurgitation (mitral [MR], tricuspid [TR], and aortic [AR]), qualitative biventricular size and function, presence/size of pericardial effusion, LV shortening fraction (LVSF), and LVEF based on Simpson’s biplane or two-dimensional analysis from an apical four chamber image only, if apical two chamber imaging was inadequate. Conventional measurements of diastolic function were obtained, including mitral valve pulsed-wave Doppler peak early (E) and late (A) diastolic inflow velocities as well as the peak early (e’) velocity of the septal and lateral aspect of the mitral valve annulus using tissue Doppler imaging. The averaged septal and lateral e’ measurements were used for calculation of the E/e’ ratio.

### Strain Analysis

Using 2D speckle-tracking software (TOMTEC-ARENA TTA2.42.00, TOMTEC, Unterschleissheim, Germany), myocardial deformational analysis was performed offline using digitally stored images from each admission echocardiogram. Optimal apical 4C images were chosen for measurement of phasic LAS, LV longitudinal strain (LVLS), and right ventricular longitudinal strain (RVLS) where the entire chamber of interest was visualized throughout the cardiac cycle.

LAS parameters were obtained by automatic software tracking of the LA endomyocardial border at end-systole and end-diastole with small, incremental manual adjustment, if needed, to ensure exclusion of the left atrial appendage and pulmonary veins. Zero strain reference was set at LV end-diastole.^32^ The LAS curve generated by the software over a single cardiac cycle was reviewed and the individual functional components (LAS-r, LAS-cd, and LAS-ct) were recorded. If a LAS-ct peak was not identifiable, the absence of this sub-component was recorded.

RVLS was obtained using the same software, with automated tracing of the RV endomyocardial border and manual adjustments from an apical or RV-focused 4C view, to generate a strain curve over a single cardiac cycle. LVLS was obtained using an apical 4C view with similar automated tracing of the endomyocardial border and manual adjustment. In cases with suboptimal visualization of a cardiac chamber of interest, the corresponding strain measurement was not obtained.

### Inter- and Intra-Rater Reliability

For inter-rater reliability assessment of LAS, primary measurements were made by one author (BJ) and a random subset (n=45) was repeated by a second observer (EB). Of this subset, ten patients did not have an identifiable LAS-ct peak resulting in fewer patients (n=35) for LAS-cd and LAS-ct reliability testing. For the same subset, the primary observer (BJ) repeated measurements two months later for intra-rater reliability assessment.

### Statistics

Demographics, clinical characteristics, and echocardiographic parameters were summarized using median and interquartile range (IQR). For categorical variables, counts and percentages were reported. The overall MIS-C cohort and controls were compared using a nonparametric Wilcoxon rank sum test for distribution skewed continuous variables and chi-squared or Fisher’s exact test for categorical variables. The MIS-C subgroups of cardiac injury and no cardiac injury were similarly compared. Correlation analysis was performed to measure the linear relationship between admission laboratory values and each strain parameter, assessed by Pearson correlation coefficient (r) with 95% confidence intervals (CI). Strength of each correlation was defined as strong (|r| >0.5), moderate (0.3< |r| ≤ 0.5), or weak (|r|<0.3). Association analysis of echocardiographic parameters and the outcome of cardiac injury was performed in univariable logistic regressions. Using only cases with complete data, a multivariable regression model was fitted to evaluate the association between cardiac injury and risk factors that were significant at p<0.1 in univariable analysis, with adjustments based upon clinical reasoning to minimize multicollinearity. Odds ratio (OR) and 95% CI were reported. Multicollinearity was assessed by generalized variance inflation factor (VIF) and a cutoff at 2.24, which is equivalent to VIF<5.^33^ Inter- and intra-rater reliability were assessed by intra-class correlation coefficients (ICC) and 95% CI. The lower bound of the 95% CI was used to interpret levels of agreement with 0.5-0.7, 0.75-0.9 and >0.9 indicating fair, good, and excellent agreements, respectively.^34^ Statistical significance was assessed at the p<0.05 level. Statistical analyses were implemented using R version 4.1.2.^35^

## Results

### Patient Characteristics

Of the 118 patients who met the MIS-C inclusion criteria, 59 (50%) had evidence of cardiac injury at the time of admission. The study group and controls were similar except for race, with higher proportions of Black or African American and Native Hawaiian or Pacific Islander in the control group (Table 1). Compared to MIS-C patients without cardiac injury, those with cardiac injury were older, had a higher body mass index (BMI), more frequently admitted to the ICU, treated with higher levels of respiratory support, and had higher inflammatory laboratory values (Table 1). One MIS-C patient in the cardiac injury subgroup underwent ECMO support.

**Table 1.**
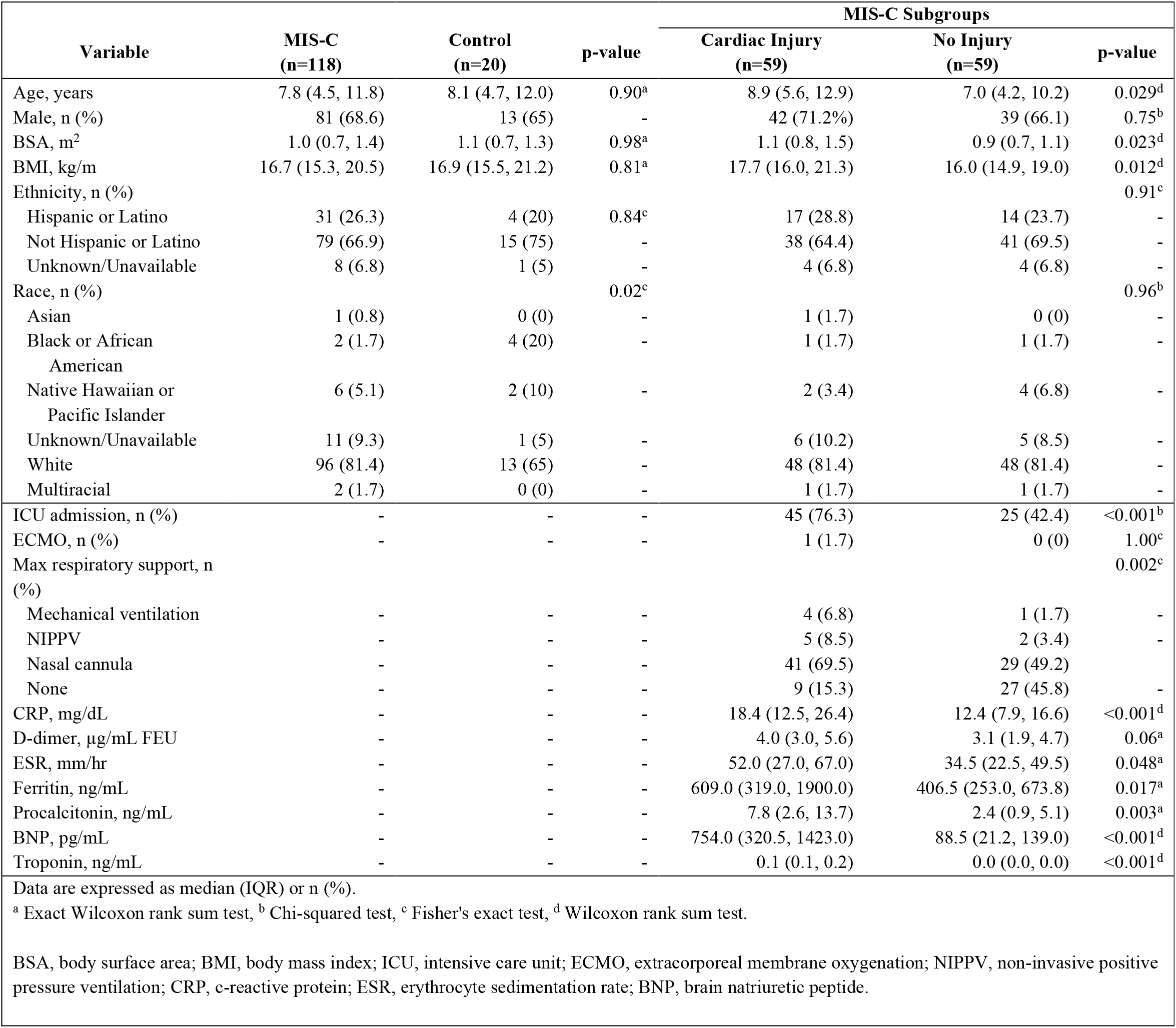
Clinical characteristics of the MIS-C study population, controls, and MIS-C subgroups with and without cardiac injury.

### Echocardiographic Findings

Of the study group, echocardiographic imaging was adequate for strain analysis in 113 (96%) for LAS, 110 (93%) for LVLS, and 91 (77%) for RVLS. Echocardiographic abnormalities in the MIS-C study group (Table 2) included TR greater than trace (17.8%), MR greater than trace (18.6%), RV dilation (2.5%), RV systolic dysfunction (8.5%), LV dilation (4.2%), LV systolic dysfunction (15.3%), and the presence of a small pericardial effusion (7.6%). No MIS-C patients had AR greater than trace, a pericardial effusion greater than small, or echocardiographic evidence of tamponade physiology. Compared to controls, MIS-C patients had lower LVEF and LVSF, and less negative LVLS and RVLS (Table 2, Figure 1). Of the conventional diastolic indices, only E/A and averaged E/e’ differed between MIS-C and controls. Compared to controls, MIS-C patients had reduced LAS-r (31.8 vs. 43.1%, p<0.001) and less negative LAS-cd (−28.8 vs. -34.5%, p=0.006) and LAS-ct (−5.2 vs. -9.3%, p<0.001). No LAS-ct peak was identified in n=65 (55%) MIS-C patients but was present in all controls (p<0.001).

**Table 2.**
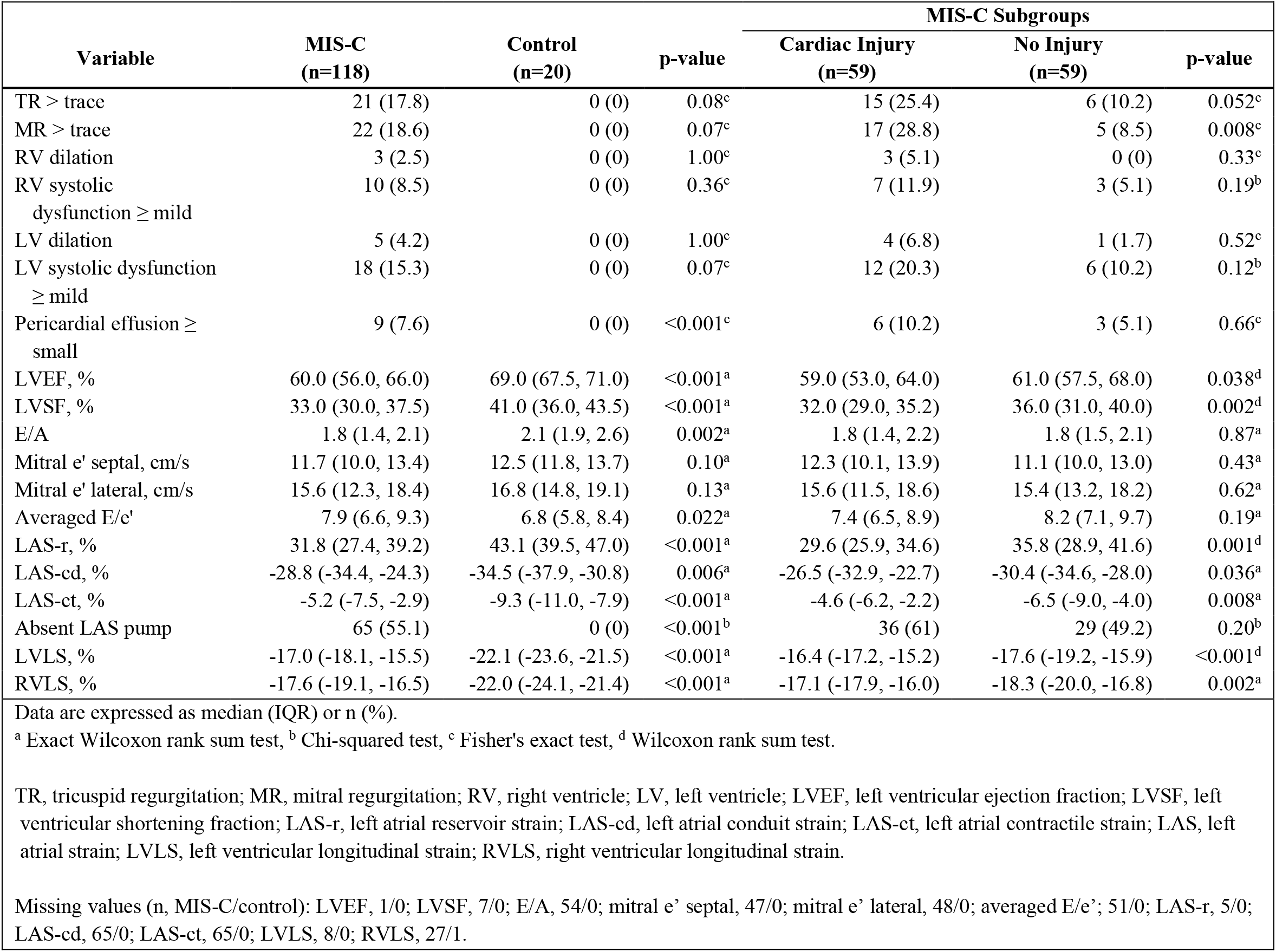
Echocardiographic parameters of the MIS-C study population, controls, and MIS-C subgroups with and without cardiac injury.

**Figure 1.**
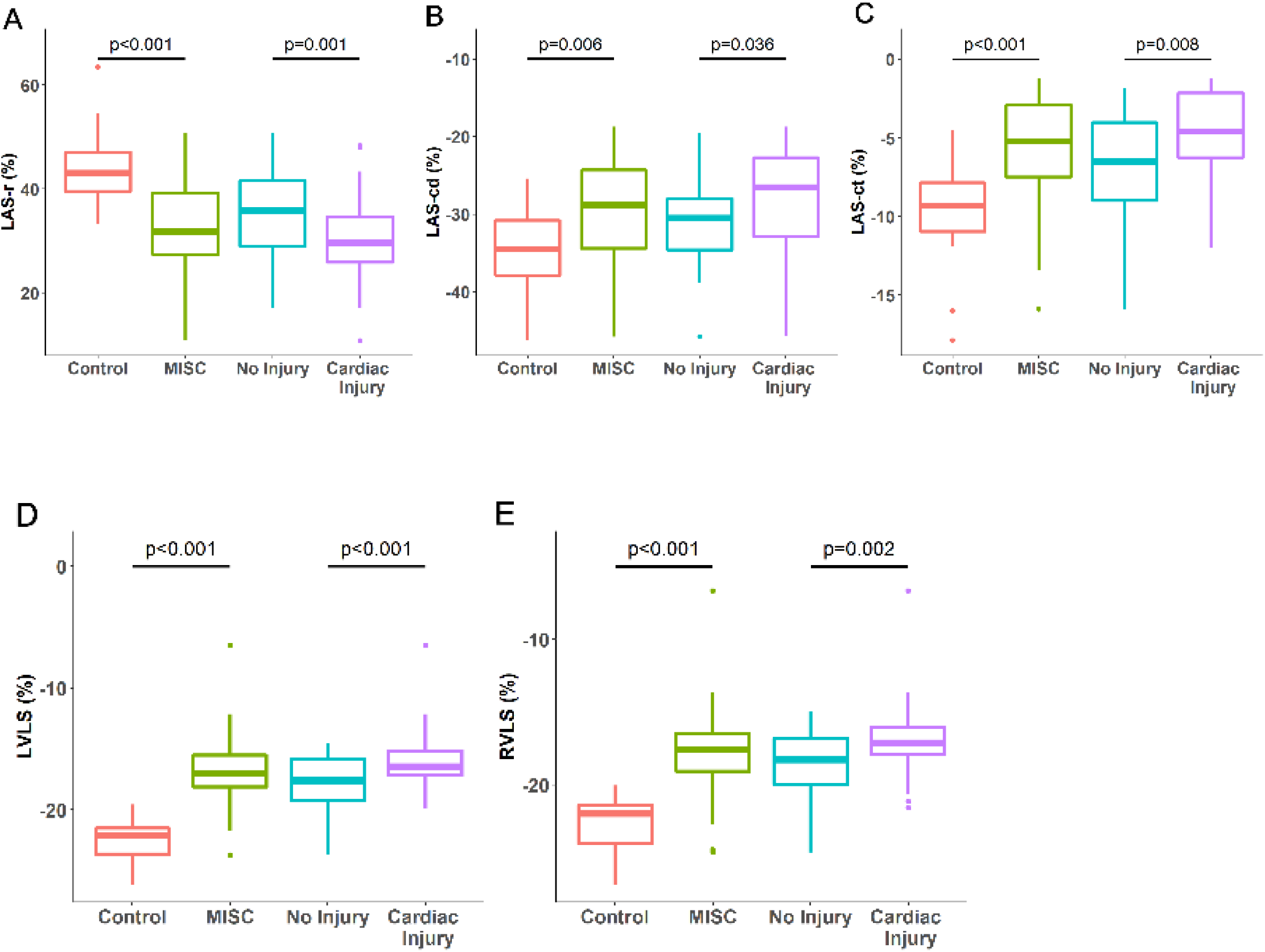
Comparison of strain parameters between MIS-C study population vs. controls and between MIS-C subgroups with vs. without cardiac injury.

A higher proportion of MIS-C patients with cardiac injury had at least mild MR compared to patients with no cardiac injury (28.8 vs. 8.5%, p=0.008). Median values for LVEF, LVSF, LVLS, and RVLS were reduced for those with cardiac injury compared to those without injury. Compared to the MIS-C subgroup with no cardiac injury, the cardiac injury subgroup had reduced LAS-r (29.6 vs. 35.8%, p=0.001) and less negative LAS-cd (−26.5 vs. -30.4%, p=0.036) and LAS-ct (−4.6 vs. -6.5%, p=0.008). Absence of an identifiable LAS-ct peak was similar between MIS-C subgroups (cardiac injury subgroup, 61% vs. no cardiac injury subgroup, 49%; p=0.2).

### Correlation Between Echocardiographic Parameters and Admission Labs

Admission D-dimer and ferritin did not correlate with any echocardiographic parameters. CRP correlated weakly with averaged E/e’ (r=0.29, p=0.023) and LAS-r (r=-0.2, p=0.039; Table 3). ESR correlated moderately with LAS-ct (r=-0.41, p=0.007). For inflammatory markers, the strongest correlation existed between procalcitonin and averaged E/e’ (r=0.55, p=0.001) and RVLS (r=0.54, p<0.001).

**Table 3.** Pearson correlation analysis of echocardiographic and laboratory values at admission for the MIS-C study population.

| Variable | CRP | ESR | Procalcitonin | BNP | Troponin |
| --- | --- | --- | --- | --- | --- |
| LVEF | -0.13 (-0.31, 0.06) | 0.11 (-0.1, 0.31) | **-0.38 (-0.57, -0.15)** | **-0.26 (-0.42, -0.08)** | **-0.29 (-0.45, -0.11)** |
| LVSF | -0.17 (-0.35, 0.02) | 0.11 (-0.1, 0.31) | **-0.33 (-0.53, -0.09)** | **-0.34 (-0.5, -0.16) \*** | **-0.26 (-0.42, -0.07)** |
| E/A | 0 (-0.25, 0.25) | 0.14 (-0.13, 0.39) | -0.25 (-0.52, 0.07) | -0.15 (-0.38, 0.1) | 0.19 (-0.06, 0.42) |
| Mitral e' septal | -0.02 (-0.26, 0.22) | -0.17 (-0.41, 0.1) | **-0.43 (-0.67, -0.11)** | -0.15 (-0.37, 0.09) | 0.24 (0, 0.45) |
| Mitral e' lateral | -0.05 (-0.29, 0.19) | -0.06 (-0.32, 0.21) | -0.03 (-0.37, 0.32) | -0.03 (-0.27, 0.21) | 0.09 (-0.15, 0.32) |
| Averaged E/e' | **0.29 (0.04, 0.5)** | 0.17 (-0.1, 0.42) | **0.55 (0.25, 0.76)** | 0.17 (-0.08, 0.39) | -0.12 (-0.35, 0.13) |
| LAS-r | **-0.2 (-0.37, -0.01)** | 0.12 (-0.08, 0.32) | -0.25 (-0.46, 0) | **-0.39 (-0.54, -0.22) \*** | **-0.21 (-0.38, -0.03)** |
| LAS-cd | 0.12 (-0.16, 0.39) | 0.07 (-0.24, 0.37) | **0.34 (-0.01, 0.61)** | 0.18 (-0.09, 0.43) | **0.28 (0.01, 0.51)** |
| LAS-ct | -0.03 (-0.31, 0.25) | **-0.41 (-0.63, -0.12)** | 0.28 (-0.07, 0.57) | **0.31 (0.04, 0.54)** | 0.12 (-0.15, 0.38) |
| LVLS | 0.13 (-0.06, 0.32) | -0.04 (-0.24, 0.17) | **0.35 (0.11, 0.55)** | **0.31 (0.13, 0.47) \*** | **0.24 (0.05, 0.41)** |
| RVLS | 0.16 (-0.06, 0.36) | -0.02 (-0.25, 0.21) | **0.54 (0.29, 0.72) \*** | **0.27 (0.07, 0.45)** | 0.13 (-0.08, 0.33) |
| Data expressed as Pearson correlation coefficient (95% CI). |  |  |  |  |  |
| Bold font indicates p <0.05 and * indicates p <0.001. Ferritin and D-dimer are not displayed since no variables correlated with p <0.05. |  |  |  |  |  |
| LVEF, left ventricular ejection fraction; LVSF, left ventricular shortening fraction; LAS-r, left atrial reservoir strain; LAS-cd, left atrial conduit strain; LAS-ct, left atrial contractile strain; LVLS, left ventricular longitudinal strain; RVLS, right ventricular longitudinal strain. |  |  |  |  |  |

BNP had a moderately strong correlation (Figure 2) with LAS-r (r=-0.39, p<0.001), LVSF (r=-0.34, p<0.001), LAS-ct (r=0.31, p=0.023), and LVLS (r=0.31, p<0.001). Troponin correlations were weak, notably with LVEF (r=-0.29, p=0.002), LAS-cd (r=0.28, p=0.041), and LVSF (r=-0.26, p=0.007).

**Figure 2.**
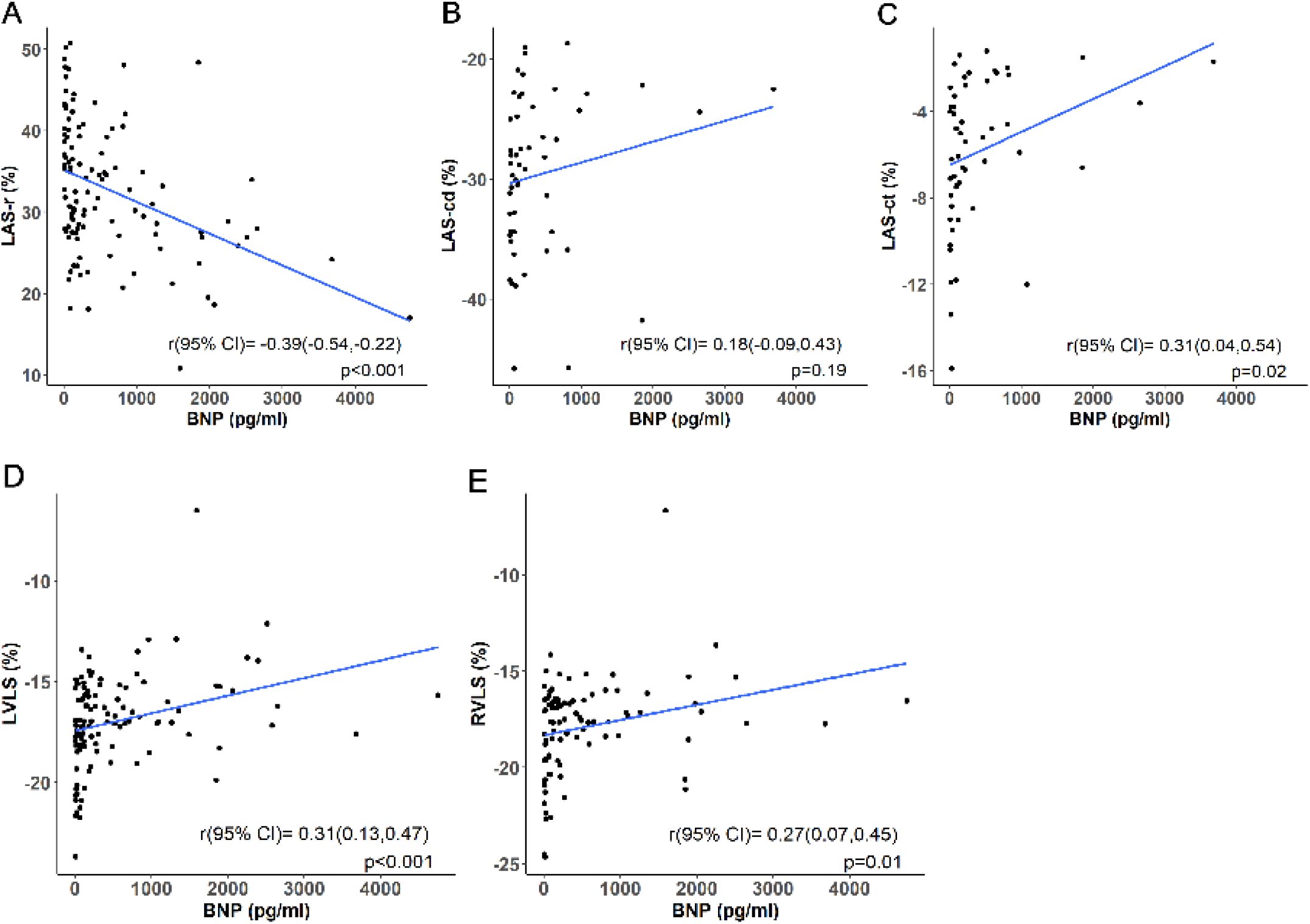
Correlation analysis for echocardiographic strain parameters and admission BNP in the MIS-C study population.

### Univariable and Multivariable Logistic Regression

In univariable analysis (Table 4), cardiac injury was associated with LVSF (OR 0.91, 95% CI 0.86-0.97, p=0.005, LAS-r (OR 0.92, 95% CI 0.88-0.97, p=0.002), LAS-ct (OR 1.28, 95% CI 1.06-1.6, p=0.016), LVLS (OR 1.45, 95% CI 1.22-1.91, p<0.001), and RVLS (OR 1.45, 95% CI 1.17-1.88, p=0.002). A multivariable logistic regression model, which included LAS-r, LVLS, and RVLS (LAS-ct and LVEF were dropped due to missingness or multicollinearity), did not identify any variables independently associated with cardiac injury.

**Table 4.** Univariable and multivariable logistic regression analyses of echocardiographic parameter association with cardiac injury in the MIS-C study population.

| Variable | Univariable |  | Multivariable |  |
| --- | --- | --- | --- | --- |
|  | OR (95% CI) | p-value | OR (95% CI) | p-value |
| LVEF | 0.96 (0.92,1) | 0.06 | - | - |
| LVSF | 0.91 (0.86,0.97) | 0.005 | - | - |
| E/A | 0.75 (0.29,1.87) | 0.54 | - | - |
| Mitral e' septal | 1.05 (0.89,1.24) | 0.59 | - | - |
| Mitral e' lateral | 0.97 (0.88,1.08) | 0.62 | - | - |
| Averaged E/e' | 0.94 (0.78,1.1) | 0.44 | - | - |
| LAS-r | 0.92 (0.88,0.97) | 0.002 | 0.96 (0.9,1.03) | 0.27 |
| LAS-cd | 1.08 (0.99,1.19) | 0.10 | - | - |
| LAS-ct | 1.28 (1.06,1.61) | 0.016 | - | - |
| LVLS | 1.5 (1.22,1.91) | <0.001 | 1.19 (0.83,1.72) | 0.35 |
| RVLS | 1.45 (1.17,1.88) | 0.002 | 1.2 (0.87,1.7) | 0.28 |
| Multivariable model based upon n=88 with no evidence of multicollinearity between independent variables. |  |  |  |  |
| LVEF, left ventricular ejection fraction; LVSF, left ventricular shortening fraction; LAS-r, left atrial reservoir strain; LAS-cd, left atrial conduit strain; LAS-ct, left atrial contractile strain; LVLS, left ventricular longitudinal strain; RVLS, right ventricular longitudinal strain. |  |  |  |  |

### Inter- and Intra-Rater Reliability

Inter-rater reliability was good to excellent for LAS-r, good for LVLS and RVLS, and fair for LAS-cd and LAS-ct (Table 5). Intra-rater reliability was good for LAS-r and LAS-cd, and fair for LAS-ct.

**Table 5.** Inter- and intra-rater reliability testing for LAS analysis.

| **Variable** | **Inter-rater** |  | **Intra-rater** |  | **n** |
| --- | --- | --- | --- | --- | --- |
|  | **ICC** | **95% CI** | **ICC** | **95% CI** |  |
| LAS-r | 0.95 | (0.9 ,0.97) | 0.87 | (0.78 ,0.93) | 45 |
| LAS-cd | 0.81 | (0.63 ,0.9) | 0.92 | (0.85 ,0.96) | 35 |
| LAS-ct | 0.76 | (0.56 ,0.87) | 0.86 | (0.74 ,0.93) | 35 |
| ICC, intra-class correlation coefficient, LAS-r, left atrial reservoir strain; LAS-cd, left atrial conduit strain; LAS-ct, left atrial contractile strain. |  |  |  |  |  |

## Discussion

Our study confirmed the reproducibility of phasic LAS analysis in children with MIS-C while adding novel insight into the global cardiac dysfunction previously described in this population.^11-18^ Phasic LAS components differed between children with MIS-C and healthy controls, and more specifically, between MIS-C patients with and without cardiac injury, even when standard echocardiographic indices were non-discriminatory. Of the phasic LAS components, LAS-r had the strongest correlation with admission BNP, although whether this reflects atrial dysfunction versus the degree of volume resuscitation remains unclear. Our findings otherwise demonstrated limited associations between admission echocardiographic parameters and laboratory markers except for procalcitonin, which had a strong association with averaged E/e’ and RVLS. The strength of these associations with procalcitonin, an inflammatory marker known to be elevated in MIS-C as well as in bacterial and severe viral infections,^36^ versus other inflammatory markers remains incompletely understood. We found that neither conventional diastolic indices nor LVEF correlated with BNP or troponin levels. Despite these findings, no echocardiographic strain parameters were independently associated with cardiac injury as we defined it in our study.

Consistent with prior studies, we found LAS-r was a highly reproducible measurement with more variability in LAS-cd and LAS-ct measurements between observers.^37,38^ Although previous reports have generally included only LAS-r in LAS analysis, LA function is complex and incompletely represented by a singular value. Prior qualitative analyses of LA strain curves in children with MIS-C identified general curve flattening and the absence of a discernable LAS-ct peak.^16,17^ While absence of an identifiable LAS-ct peak occurred in over half of our study group, we notably did not observe this phenomenon in any control patients. Although this metric performed less well in differentiating MIS-C patients with and without cardiac injury, the binary presence or absence of an identifiable LAS-ct peak may be a useful metric for identification of LA dysfunction in patients with MIS-C.

LAS deviations and cardiac biomarker elevation in children with MIS-C have suggested some degree of diastolic dysfunction, which was not identifiable by conventional echocardiographic indices. In our study, conventional diastolic indices remained within normal adult ranges, despite small differences in E/A and averaged E/e’ between groups.^39^ Since adult-derived cutoffs have not been generalizable to pediatric populations due to inadequate discriminatory ability,^40^ phasic LAS analysis may fill this clinical gap.

Our phasic LAS data may be the starting point for understanding the natural history of diastolic dysfunction in MIS-C. Previously described categorial gradations of left ventricular diastolic dysfunction using phasic LAS analysis may be a useful framework for interpreting our results. Ta *et al*. (2020) described a binary classification of compensatory versus decompensatory LA dysfunction based upon pressure or volume loading in patients with congenital heart disease.^26^ In contrast to early diastolic dysfunction, in which LAS-r and LAS-cd decrease with a compensatory increase in LAS-ct, decompensatory LA dysfunction was defined as a decrease in all three parameters. We found a decrease in all three LAS components in MIS-C patients versus controls and as well as in MIS-C patients with versus without cardiac injury, suggesting a later decompensatory stage of LA dysfunction at admission as outlined by Ta *et al*. (2020) or a similar grade 2 diastolic dysfunction as described by Singh *et al*. (2017).^20,26^

Limitations of our investigation include the single center and retrospective nature of the study. We analyzed admission echocardiograms, although patients with MIS-C present for care at varying timepoints and their echocardiographic parameters may be impacted by disease evolution. In addition, by defining cardiac injury using admission laboratory values, our MIS-C cardiac injury subgroup did not include patients who developed elevated BNP and/or troponin values later in the hospital course. Despite these limitations, the reproducibility of phasic LAS analysis increases the likelihood of its use in future research and clinical applications. The establishment of normal pediatric values for phasic LAS components will be essential for its transition from a research tool into clinical practice.

## Conclusion

LAS analysis in children with MIS-C was reproducible and may identify diastolic dysfunction not discernable by conventional echocardiographic indices. Although phasic LAS components were more highly associated with cardiac injury than conventional echocardiographic parameters, no strain parameters were independently associated with cardiac injury at the time of admission in MIS-C. Further longitudinal investigation of LAS will improve our understanding of diastolic dysfunction within the natural history of MIS-C.

## Data Availability

All data produced in the present study are available upon reasonable request to the authors.

## Acknowledgments

This investigation was supported by the University of Utah Study Design and Biostatistics Center, with funding in part from the National Center for Research Resources and the National Center for Advancing Translational Sciences, National Institutes of Health, through Grant 8UL1TR000105 (formerly UL1RR025764).

## Abbreviations

LAS: Left atrial strain
LAS-r: Left atrial reservoir strain
LAS-cd: Left atrial conduit strain
LAS-ct: Left atrial contractile strain
LVLS: Left ventricular longitudinal strain
RVLS: Right ventricular longitudinal strain

## Notes

### Competing Interest Statement

The authors have declared no competing interest.

### Funding Statement

This study was funded in part by the National Center for Research Resources and the National Center for Advancing Translational Sciences, National Institutes of Health, through Grant 8UL1TR000105 (formerly UL1RR025764).

### Author Declarations

The Institutional Review Board of the University of Utah and the Intermountain Healthcare Privacy Board gave ethical approval for this work.

## References

1. Verdoni L, Mazza A, Gervasoni A, et al. An outbreak of severe Kawasaki-like disease at the Italian epicentre of the SARS-CoV-2 epidemic: an observational cohort study. Lancet. 2020;395(10239):1771–1778. doi: 10.1016/S0140-6736(20)31103-X

2. Dufort EM, Koumans EH, Chow EJ, et al. Multisystem Inflammatory Syndrome in Children in New York State. N Engl J Med. 2020;383(4):347–358. doi: 10.1056/NEJMoa2021756

3. Feldstein LR, Rose EB, Horwitz SM, et al. Multisystem Inflammatory Syndrome in U.S. Children and Adolescents. N Engl J Med. 2020;383(4):334–346. doi:10.1056/NEJMoa2021680

4. Feldstein LR, Tenforde MW, Friedman KG, et al. Characteristics and Outcomes of US Children and Adolescents With Multisystem Inflammatory Syndrome in Children (MIS-C) Compared With Severe Acute COVID-19. JAMA. 2021;325(11):1074–1087. doi:10.1001/jama.2021.2091

5. Valverde I, Singh Y, Sanchez-de-Toledo J, et al. Acute Cardiovascular Manifestations in 286 Children With Multisystem Inflammatory Syndrome Associated With COVID-19 Infection in Europe. Circulation. 2021;143(1):21–32. doi:10.1161/CIRCULATIONAHA.120.050065

6. Capone CA, Subramony A, Sweberg T, et al. Characteristics, Cardiac Involvement, and Outcomes of Multisystem Inflammatory Syndrome of Childhood Associated with severe acute respiratory syndrome coronavirus 2 Infection. J Pediatr. 2020;224:141–145. doi:10.1016/j.jpeds.2020.06.044

7. Riphagen S, Gomez X, Gonzalez-Martinez C, et al. Hyperinflammatory shock in children during COVID-19 pandemic. Lancet. 2020;395(10237):1607–1608. doi:10.1016/S0140-6736(20)31094-1

8. Whittaker E, Bamford A, Kenny J, et al. Clinical Characteristics of 58 Children With a Pediatric Inflammatory Multisystem Syndrome Temporally Associated With SARS-CoV-2. JAMA. 2020;324(3):259–269. doi:10.1001/jama.2020.10369

9. Friedman KG, Harrild DM, Newburger JW. Cardiac Dysfunction in Multisystem Inflammatory Syndrome in Children: A Call to Action. J Am Coll Cardiol. 2020;76(17):1962–1964. doi:10.1016/j.jacc.2020.09.002

10. Belay ED, Abrams J, Oster ME, et al. Trends in Geographic and Temporal Distribution of US Children With Multisystem Inflammatory Syndrome During the COVID-19 Pandemic. JAMA Pediatr. 2021;175(8):837–845. doi:10.1001/jamapediatrics.2021.0630

11. Gaitonde M, Ziebell D, Kelleman MS, et al. COVID-19-Related Multisystem Inflammatory Syndrome in Children Affects Left Ventricular Function and Global Strain Compared with Kawasaki Disease. J Am Soc Echocardiogr. 2020;33(10):1285–1287. doi:10.1016/j.echo.2020.07.019

12. Kobayashi R, Dionne A, Ferraro A, et al. Detailed Assessment of Left Ventricular Function in Multisystem Inflammatory Syndrome in Children, Using Strain Analysis. CJC Open. 2021;3(7):880–887. doi:10.1016/j.cjco.2021.02.012

13. He M, Leone DM, Frye R, et al. Longitudinal Assessment of Global and Regional Left Ventricular Strain in Patients with Multisystem Inflammatory Syndrome in Children (MIS-C). Pediatr Cardiol. 2022;43(4):844–854. doi:10.1007/s00246-021-02796-7

14. Basu S, Kim EJ, Sharron MP, et al. Strain Echocardiography and Myocardial Dysfunction in Critically Ill Children With Multisystem Inflammatory Syndrome Unrecognized by Conventional Echocardiography: A Retrospective Cohort Analysis. Pediatr Crit Care Med. 2022;23(3):e145–e152. doi:10.1097/PCC.0000000000002850

15. Leal GN, Astley C, Lima MS, et al. Segmental cardiac strain assessment by two-dimensional speckle-tracking echocardiography in surviving MIS-c patients: Correlations with myocardial flow reserve (MFR) by 13 N-ammonia PET-CT. Microcirculation. 2022;29(3):e12750. doi:10.1111/micc.12750

16. Matsubara D, Kauffman HL, Wang Y, et al. Echocardiographic Findings in Pediatric Multisystem Inflammatory Syndrome Associated With COVID-19 in the United States. J Am Coll Cardiol. 2020;76(17):1947–1961. doi:10.1016/j.jacc.2020.08.056

17. Matsubara D, Chang J, Kauffman HL, et al. Longitudinal Assessment of Cardiac Outcomes of Multisystem Inflammatory Syndrome in Children Associated With COVID-19 Infections. J Am Heart Assoc. 2022;11(3):e023251. doi:10.1161/JAHA.121.023251

18. Sanil Y, Misra A, Safa R, et al. Echocardiographic Indicators Associated with Adverse Clinical Course and Cardiac Sequelae in Multisystem Inflammatory Syndrome in Children with Coronavirus Disease 2019. J Am Soc Echocardiogr. 2021;34(8):862–876. doi:10.1016/j.echo.2021.04.018

19. Kavurt AV, Bağrul D, Gül AEK, et al. Echocardiographic Findings and Correlation with Laboratory Values in Multisystem Inflammatory Syndrome in Children (MIS-C) Associated with COVID-19. Pediatr Cardiol. 2022;43(2):413–425. doi:10.1007/s00246-021-02738-3

20. Singh A, Addetia K, Maffessanti F, et al. LA Strain for Categorization of LV Diastolic Dysfunction. JACC Cardiovasc Imaging. 2017;10(7):735–743. doi:10.1016/j.jcmg.2016.08.014

21. Thomas L, Marwick TH, Popescu BA, et al. Left Atrial Structure and Function, and Left Ventricular Diastolic Dysfunction: JACC State-of-the-Art Review. J Am Coll Cardiol. 2019;73(15):1961–1977. doi:10.1016/j.jacc.2019.01.059

22. Singh A, Medvedofsky D, Mediratta A, et al. Peak left atrial strain as a single measure for the non-invasive assessment of left ventricular filling pressures. Int J Cardiovasc Imaging. 2019;35(1):23–32. doi:10.1007/s10554-018-1425-y

23. Meel R, Khandheria BK, Peters F, et al. Effects of age on left atrial volume and strain parameters using echocardiography in a normal black population. Echo Res Pract. 2016;3(4):115–123. doi:10.1530/ERP-16-0038

24. Yoshida K, Obokata M, Kurosawa K, et al. Effect of Sex Differences on the Association Between Stroke Risk and Left Atrial Anatomy or Mechanics in Patients With Atrial Fibrillation. Circ Cardiovasc Imaging. 2016;9(10):e004999. doi:10.1161/CIRCIMAGING.116.004999

25. Liao JN, Chao TF, Kuo JY, et al. Age, Sex, and Blood Pressure-Related Influences on Reference Values of Left Atrial Deformation and Mechanics From a Large-Scale Asian Population. Circ Cardiovasc Imaging. 2017;10(10):e006077. doi:10.1161/CIRCIMAGING.116.006077

26. Ta HT, Alsaied T, Steele JM, et al. Atrial Function and Its Role in the Non-invasive Evaluation of Diastolic Function in Congenital Heart Disease. Pediatr Cardiol. 2020;41(4):654–668. doi:10.1007/s00246-020-02351-w

27. Centers for Disease Control and Prevention. Emergency preparedness and response: multisystem inflammatory syn drome in children (MIS-C) associated with coronavirus disease 2019 (COVID-19). Health advisory (https://emergency.cdc.gov/han/2020/han00432.asp)

28. Harris PA, Taylor R, Thielke R, et al. Research electronic data capture (REDCap)--a metadata-driven methodology and workflow process for providing translational research informatics support. J Biomed Inform. 2009;42(2):377–381. doi:10.1016/j.jbi.2008.08.010

29. Harris PA, Taylor R, Minor BL, et al. The REDCap consortium: Building an international community of software platform partners. J Biomed Inform. 2019;95:103208. doi:10.1016/j.jbi.2019.103208

30. Lai WW, Geva T, Shirali GS, et al. Guidelines and standards for performance of a pediatric echocardiogram: a report from the Task Force of the Pediatric Council of the American Society of Echocardiography. J Am Soc Echocardiogr. 2006;19(12):1413–1430. doi:10.1016/j.echo.2006.09.001

31. Lopez L, Colan SD, Frommelt PC, et al. Recommendations for quantification methods during the performance of a pediatric echocardiogram: a report from the Pediatric Measurements Writing Group of the American Society of Echocardiography Pediatric and Congenital Heart Disease Council. J Am Soc Echocardiogr. 2010;23(5):465–577. doi:10.1016/j.echo.2010.03.019

32. Badano LP, Kolias TJ, Muraru D, et al. Standardization of left atrial, right ventricular, and right atrial deformation imaging using two-dimensional speckle tracking echocardiography: a consensus document of the EACVI/ASE/Industry Task Force to standardize deformation imaging [published correction appears in Eur Heart J Cardiovasc Imaging. 2018 Jul 1;19(7):830-833]. Eur Heart J Cardiovasc Imaging. 2018;19(6):591–600. doi:10.1093/ehjci/jey042

33. James, Gareth, Daniela Witten, Trevor Hastie, and Robert Tibshirani. 2014. An Introduction to Statistical Learning: With Applications in R. Springer Publishing Company, Incorporated.

34. Koo TK, Li MY. A Guideline of Selecting and Reporting Intraclass Correlation Coefficients for Reliability Research [published correction appears in J Chiropr Med. 2017 Dec;16(4):346]. J Chiropr Med. 2016;15(2):155–163. doi:10.1016/j.jcm.2016.02.012

35. R Core Team (2020). R: A language and environment for statistical computing. R Foundation for Statistical Computing, Vienna, Austria. URL https://www.R-project.org/.)

36. Roberts JE, Campbell JI, Gauvreau K, et al. Differentiating multisystem inflammatory syndrome in children: a single-centre retrospective cohort study. Arch Dis Child. 2022 Mar;107(3):e3. doi:10.1136/archdischild-2021-322290

37. Hope KD, Wang Y, Banerjee MM, et al. Left atrial mechanics in children: insights from new applications of strain imaging. Int J Cardiovasc Imaging. 2019;35(1):57–65. doi:10.1007/s10554-018-1429-7

38. Rausch K, Shiino K, Putrino A, et al. Reproducibility of global left atrial strain and strain rate between novice and expert using multi-vendor analysis software. Int J Cardiovasc Imaging. 2019;35(3):419–426. doi:10.1007/s10554-018-1453-7

39. Nagueh SF, Smiseth OA, Appleton CP, et al. Recommendations for the Evaluation of Left Ventricular Diastolic Function by Echocardiography: An Update from the American Society of Echocardiography and the European Association of Cardiovascular Imaging. J Am Soc Echocardiogr. 2016 Apr;29(4):277–314. doi:10.1016/j.echo.2016.01.011

40. Dragulescu A, Mertens L, Friedberg MK. Interpretation of left ventricular diastolic dysfunction in children with cardiomyopathy by echocardiography: problems and limitations. Circ Cardiovasc Imaging. 2013;6(2):254–261. doi:10.1161/CIRCIMAGING.112.000175

